# Why sovereignty in PPR vaccination matters for pastoral women: Exploring experiences and perspectives in a qualitative study in the Somali region, Ethiopia

**DOI:** 10.1101/2024.09.18.24313383

**Authors:** Valerie Hungerbühler, Buckary Barkadle, Amina Mohamed Hussein, Sofia C. Zambrano, Mohammed Digale, Kebadu Simachew Belay, Flurina Derungs, Salome Dürr

## Abstract

Peste des petits ruminants (PPR) is a viral disease that significantly affects small ruminants, posing a serious threat to the livelihoods and food security of rural communities. Pastoral women are particularly impacted, as they are the primary caretakers of sheep and goats. While managing livestock can empower women, prevailing gender norms may limit this potential. This study aimed to explore women’s experiences in taking on responsibility for animal health and to analyse the challenges and opportunities presented by PPR intervention projects.

Eight focus group discussions were conducted with women from three districts in the Somali region of Ethiopia, supplemented by twelve key informant questionnaires and one key informant interview. Qualitative data were analysed using reflexive thematic analysis, while quantitative data were analysed using descriptive statistics.

The research findings present one overarching theme: ‘The importance of women’s sovereignty in PPR vaccination,’ along with two main themes. The first theme, ‘Navigating hierarchies and the feminisation of responsibility,’ comprises the subthemes ‘Women in responsibility,’ ‘Cultural norms and masculinities,’ and ‘Men losing track.’ This theme highlights women’s predominant roles in livestock management, household chores, and income generation, while it also explores traditional gender roles and the impacts of men’s khat consumption. The second main theme, ‘Knowledge privilege, dilemmas, and gaps,’ encompasses the subthemes ‘Knowledge for empowerment,’ ‘Dilemmas despite knowledge,’ and ‘Challenges due to training gaps.’ This theme explores women’s empowerment through PPR training, underscores the critical importance of unhindered access to PPR vaccines and effective cold chain maintenance, addresses challenges such as vaccine hesitancy and antimicrobial usage, and stresses the need for community-based training initiatives.

The study emphasises a shift from the traditional gendered division of labor toward an increased burden of responsibility on women, a trend that appears to have been reinforced by anti-poverty programs and further intensified by men’s khat consumption. Training in animal health, particularly in PPR vaccination, has been identified as an area where women are eager to engage. To support this, the study recommends adopting ‘train-the-trainer’ and community-based approaches, while ensuring access to resources. These strategies must be gender-responsive to empower women in PPR management and reduce their workloads in the long term.

**One health impact statement:** This research highlights the importance of involving pastoral women in managing Peste des petits ruminants (PPR), a viral disease that significantly affects sheep and goats. It also emphasises the risk of overburdening women in poverty reduction efforts and stresses the need for gender-responsive initiatives to reduce their workload in the long term.

By empowering women in animal health management through such initiatives, both animal health and the livelihoods and food security of rural communities are improved. This underscores the value of a One Health approach, which integrates human, veterinary, and social sciences to comprehensively address health challenges.

The research was made possible through long-standing trust relationships between NGO staff and communities, which facilitated strong community engagement. The qualitative and reflexive approach to data analysis enabled the voices of affected individuals to be heard, allowing for the co-production of knowledge and enhancing the research’s societal relevance.

## 1. Introduction

Peste des petits ruminants (PPR), caused by a morbillivirus closely related to the rinderpest virus, is a highly contagious disease primarily affecting sheep and goats (WOAH). Its high morbidity and mortality rates pose severe socio-economic challenges, notably impacting the food security of rural communities dependent on subsistence farming (Kihu et al., 2015). The World Organisation for Animal Health (WOAH) and the Food and Agriculture Organization (FAO) are pursuing a global strategy to eradicate the disease by 2030 (WOAH & FAO, 2022). This strategy draws on insights from the eradication of rinderpest and emphasises the crucial role of community animal health workers in vaccine administration and disease monitoring (WOAH). The prospects for successful eradication are promising, given the singular serotype of the disease and the absence of carrier states or sustainable reservoirs outside domestic small ruminants (WOAH). Moreover, a cost-effective vaccine conferring lifelong immunity is available (Hodgson et al., 2018). The global eradication strategy relies on the immunisation of small ruminant populations through mass vaccination campaigns (WOAH & FAO, 2022). However, these campaigns are costly and difficult to implement due to the vaccine’s thermolability, the limited accessibility and high mobility of some small ruminant populations, and the lack of precise census data and national animal identification systems for small ruminants (Fournié et al., 2018; Kumar et al., 2017). PPR remains prevalent in many low- and middle-income countries (LMICs), including Ethiopia (Dubie et al., 2022; Ejigu et al., 2023; Fournié et al., 2018), where it particularly affects pastoral women who are the primary caretakers of sheep and goats.

In many smallholder farming communities, women have significantly increased their involvement in agricultural production over the past few decades. Traditionally, women’s participation in the livestock sector varies by species. Men typically control cattle, camels, and buffalo, while women manage less profitable livestock, such as poultry and small ruminants (Hillesland et al., 2021; Hovorka, 2012). Consequently, small ruminants are particularly important to pastoral women for generating income, providing food, accumulating wealth, and conferring social status (McKune, Serra, & Touré, 2021). More recently, women have increasingly shouldered the responsibility for household survival and responded to economic opportunities in commercial agriculture (Lastarria-Cornhiel, 2006). As a result, women provide much of the work required along the livestock value chains (Omondi et al., 2022). Governmental and non-governmental organizations (NGOs) have recognised the potential of targeting women in livestock management to enhance livelihoods and reduce poverty (Chant, 2014). Evidence suggests that involving women in income allocation is more effective than involving men for improving family welfare (Adeyemi, 2010; Duflo, 2012; Duflo & Udry, 2004; Maertens & Verhofstadt, 2013; O’Brien et al., 2016; A. R. Quisumbing & Maluccio, 2003). However, intensified involvement in agriculture may disproportionately burden women’s workloads compared to men’s, adding to their caregiving and domestic responsibilities. This underscores the need to investigate and challenge existing gender norms (A. Quisumbing et al., 2023).

Gender norms are social constructs that define the roles, behaviors, and activities considered appropriate for women and men (Kariuki et al., 2022). For example, ‘restrictive masculinities’ confine men to traditional roles, thereby undermining women’s empowerment and equality, often by granting men preferential access to resources while restricting women’s access (Cislaghi et al., 2018). A 2019 report by the Organisation for Economic Cooperation and Development defines “gender-equitable masculinities” as a new form of masculinities that supports women’s empowerment and gender equality by challenging patriarchal structures (OECD, 2019).

Following this rationale, the NGOs Vétérinaires Sans Frontières Suisse and Germany (VSF-Suisse and VSF Germany) have collaboratively implemented a gender-sensitive PPR intervention project, “GIVE-Women,” to provide specialised training for women on managing small ruminant diseases, with a focus on PPR and vaccine administration, in the Somali Region of Ethiopia. The Somali Region is a distinct area in the eastern part of the country, bordering Djibouti to the north, Somalia to the east, and Kenya to the south. Somali culture is rich and diverse, deeply rooted in traditions passed down through generations (Diriye Abdullahi, 2001). The region experiences a harsh, hot, and dry climate, with sparse and erratic rainfall. Droughts and water scarcity are common challenges faced by the inhabitants (Kebede et al., 2024). Historically, many Somalis have practiced nomadic pastoralism, herding livestock across highlands and lowlands, which has increased their resilience to harsh environmental conditions (Kebede et al., 2024). While urbanisation and sedentarisation have led to lifestyle changes for some, nomadic pastoralism persists as a way of life for many communities in the region (Kebede et al., 2024). PPR is endemic in the Somali region of Ethiopia, with a 2008 study reporting a seroprevalence of 21.3% (Waret-Szkuta et al., 2008), while a more recent study in the neighboring Afar region reported a seroprevalence of 60.15 % in small ruminants (Dubie et al., 2022). In the Somali region, PPR vaccines are commonly administered during vaccination campaigns implemented by the government. Livestock keepers receive vaccinations free of charge, either in response to outbreaks or upon individual request through government animal health workers. However, according to unpublished VSF reports, such campaigns have recently become less frequent in the study area, resulting in inadequate vaccination coverage. This issue appears to be global, as Jones et al. (2016) noted that the average annual vaccination coverage reported to the WOAH by PPR-infected countries is 15%, while Singh (2011) suggests 85.4% coverage is needed to control PPR in the small ruminant population.

Various stakeholders, including governmental organisations and NGOs, are mobilising efforts to improve animal health services (FAOLEX, 2015). Notably, recognising the crucial role of women in small ruminant disease surveillance, initiatives like ‘GIVE-Women’ are emerging, all with the aim of empowering women to take on responsibilities in their communities (McKune et al., 2023; Omondi et al., 2022). On the other hand, interventions aimed at increasing women’s participation in livestock (health) management may inadvertently increase women’s workload, undermining their empowerment. Qualitative research is necessary to understand the nuances of empowerment, informing strategies for integrating women into vaccine administration, without overburdening them. This study aimed to explore women’s experiences in taking on responsibility for animal health and to analyse the challenges and opportunities presented by a PPR intervention project focused on women’s participation in vaccination efforts.

## 2. Methods

### 2.1 Study sites and community engagement

The study was conducted across three districts (Shinile, Ayshia, Kebribeyah) in Ethiopia’s Somali region, encompassing eight kebeles within these districts. A district comprises multiple kebeles, which are the smallest administrative units, similar to neighborhoods or villages. In this context, the term ‘community’ refers to the collective inhabitants of a kebele and is used to describe the affiliation of the focus group participants.

VSF-Suisse has a long history of conducting research and implementing intervention projects in the Somali region, enabling the organisation to establish long-standing and trusted relationships with project communities. These established connections facilitated effective community engagement throughout the research process.

For this study, eight communities were selected using convenience sampling. In three of these communities, some women had previously received training in animal health, including PPR vaccination, through the ‘GIVE-Women’ project conducted by VSF-Suisse (VSF-Suisse, 2024). This training is referred to as ‘PPR training’ in the following sections. Women in another three communities had received training in livestock rearing and milk hygiene. The remaining two communities had not received any prior training from VSF-Suisse.

### 2.2 Data collection

Data collection took place between April and May 2023 and consisted of eight focus group discussions (FGDs), 12 key informant (KI) questionnaires (KIQs) and one KI interview (KII). One FGD was conducted in each community, with 8–18 female participants per group. Participation was voluntary, resulting in a convenience sample of focus group participants. A female sociologist proficient in English, Arabic, Amharic, and Somali facilitated the FGDs. All sessions were voice-recorded, and detailed notes were taken throughout.

The FGDs had a two-part structure. In the first part, participants discussed their livestock assets, prevalent diseases, and experiences with PPR. This was followed by discussions on women’s experiences before and after PPR training provided by VSF-Suisse. Topics included vaccine accessibility, maintenance of the cold chain, availability of medications (“injections”), and perceptions regarding female animal health service providers. Participants also shared their concerns and recommendations for future animal health training initiatives. In the second part, we used the indicators from the Women’s Empowerment in Livestock Index (WELI) to guide discussions on social dynamics in livestock management (Galiè, Teufel, Korir, et al., 2019).

The KIQs were collected from four male experts from the pastoral development office of Jijiga, three male experts from Jijiga University, two male experts from the veterinary laboratory at Jijiga University, and three veterinary medicine master’s students of the Jijiga University, one of whom was female. These experts were selected for their knowledge of veterinary diseases, including PPR, and the animal health service system in the Somali region. The multiple-choice questionnaire included questions on the gendered division of labor in pastoral communities and women’s empowerment, along with free-text fields for respondents to provide additional comments.

Finally, the KII was conducted with a male expert from the pastoral development office (PDO), focusing specifically on the issue of khat consumption. Khat is a plant containing psychoactive substances and is commonly chewed in Ethiopia, predominantly by men, for social, religious, and stimulatory purposes.

### 2.3 Analytic process

#### 2.3.1 Qualitative data analysis

All audio data were translated and transcribed into English by the data collection team, which had a strong command of English and Somali. The transcripts underwent verification by cross-referencing the audio files and scripts with the field notes taken during the discussions. Data management involved the use of Excel, Word, and hard copies.

A reflexive approach to thematic analysis, following Braun & Clarke (2006), was employed. This involved a six-phase process: 1) dataset familiarisation, 2) rigorous and systematic coding, 3) initial theme generation, 4) theme development and review, 5) theme refining, defining, and naming, and finally 6) producing the written report. The research and analysis were grounded in an experiential approach underpinned by a critical realist ontology. The epistemological standpoint aligned with contextualism, emphasising the context-dependent nature of language and meaning. The analysis adopted an inductive approach for coding, focusing primarily on a semantic level while exploring the participants’ lived experiences. A descriptive mode of interpretation was applied, with an awareness that the researcher naturally interprets data informed by insights from theory and related research.

#### 2.3.2 Quantitative data analysis

The responses from the multiple-choice KIQs were quantitatively analysed using descriptive statistics in Excel. Additional comments from the free-text fields were included as quotes to supplement the quantitative results.

## 3. Results

### 3.1 Themes identified through reflexive analysis of FGDs

We identified ‘The importance of women’s sovereignty in PPR vaccination’ as the overarching theme encompassing the experiences and perceptions of the women who participated in the FGDs. In this context, sovereignty reflects the participants’ belief that women who received PPR training should have control, authority, and autonomy over decisions and actions related to PPR vaccination. This idea indicates that women sought empowerment to make informed decisions about vaccinating their livestock and to independently acquire and administer vaccines. Within this overarching theme, we identified two main themes: 1) ‘Navigating hierarchies and the feminisation of responsibility,’ and 2) ‘Knowledge privilege, dilemmas, and gaps,’ (Figure 1).

**Figure 1:**
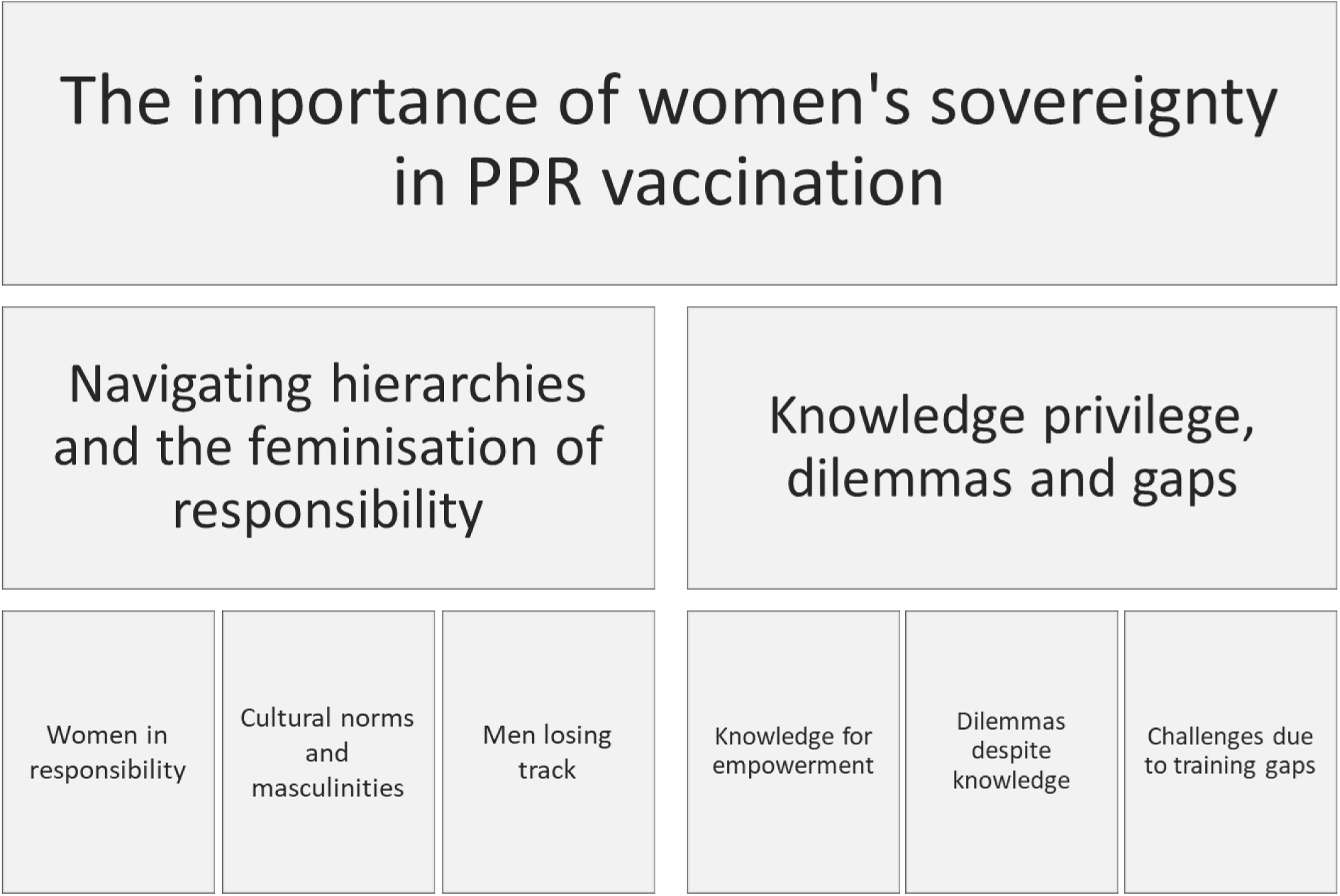
Thematic Map: Overarching theme atop, main themes in the center, subthemes below. Themes were identified during a qualitative study with women from pastoral communities in Ethiopia’s Somali region in April – May 2023.

#### 3.1.1 Navigating hierarchies and the feminisation of responsibility

The theme of ‘Navigating hierarchies and the feminisation of responsibility’ delves into a central idea with various facets, as expressed by the focus group participants. While constrained by traditional gender hierarchies, women seemed to increasingly take on additional responsibilities that extended beyond the household, where they have historically been the primary bearers of reproductive labor. Conversely, the other side of the feminisation of responsibility involved men stepping back from their duties, a trend influenced in part by khat consumption.

##### Subtheme: Women in responsibility

The first subtheme, ‘Women in responsibility,’ explores two contradictory facets. On one hand, taking on responsibility was perceived as a form of empowerment, with women reporting a sense of autonomy in certain contexts. They mentioned their responsibilities in managing livestock, accessing income-generating opportunities and credit, and controlling household expenses and purchases. Women indicated that they made certain decisions independently, and shared decision-making between men and women appeared to be common. Additionally, NGOs seemed to actively involve women in training. Women expressed a sense of power and pride, recognising their role as the backbone of the community. However, on the other hand, women constantly worked without control over their own time, dictated by the demands of life, including those of their children, husbands, and livestock.

Most participants articulated a growing sense of autonomy in livestock management, particularly regarding animal health, indicating that this independence has developed over time. Some participants noted that women undertook all household and livestock management tasks, with men shouldering merely symbolic responsibility.

> *“In livestock management, everything is in the hands of women. Men do not do anything these days.”* **A woman from Gad**
>
> “Most of the work is done by women because there are more women than men, and men are always chewing khat. Just as we take responsibility for the children, we also take responsibility for the livestock.” **A woman from Dhaga Jabis**
>
> A participant from Biyo-Kabobe articulated her perspective on the gendered division of animal health tasks with the following statement:
>
> *“Men never say, ‘You cannot do that because you are a woman.’ However, certain tasks, like camel handling, come more naturally to men. Men are stronger and can handle a jumping camel, which women may find difficult due to their clothing. When camels jump, women tend to move away, but this is not about disrespect. With tasks involving goats and sheep, women can easily manage, even with just one hand, given proper training. Sometimes, men hope women can handle camels so they are free to do nothing. There are certain jobs, like handling camels, traditionally reserved for men and even those he wants to be able to give to women.”* **A woman from Biyo-Kabobe**

Most participants indicated that decision-making regarding livestock management is no longer exclusively controlled by men, as it was in the past. Instead, shared decision-making is becoming increasingly common, with women expressing a sense of equality while still acknowledging the symbolic authority held by men.

> “*In the past, men ruled. However, we now share power, and women can decide. Nonetheless, the ultimate responsibility lies with him*.” **A woman from Guyo**

Participants reported that women can access income-generating opportunities by selling livestock and their milk. Men seemed to face challenges in earning income but recognised that women’s earnings benefit the family.

> “*We generate income from our livestock by selling the animals or their milk or khat. Occasionally, men bring in some money from relatives. All of us [women] generate income. Sometimes, we engage in group work. The husband of one of us is the only exception; he is a policeman. The income he receives from the government is brought into the family*.” **A woman from Guyo**
>
> “*Economically, women are doing much better than men, which gives us the strength to act self-determinedly*.” **A woman from Dhaga Jabis**
>
> “*My husband does not mind me selling livestock because it benefits the family*.” **A woman from Gilla**

Women appeared to have gained more control over income compared to earlier times, while men seemed to have consciously stepped back from this responsibility at times. Some participants attributed this shift to women’s stronger income management skills and a deeper understanding of their family’s needs. Men’s authority was still evident, as women often sought their approval before spending income or joining a savings group. However, men were generally supportive, recognising the benefits these actions brought to the family.

> “*In the past, it was the men, but today, women manage the income. We generate income from the livestock by selling or using animal products. We typically seek men’s consent or inform them, but we manage the income. I must get my husband’s approval, then I can do what I want. They give to the women whatever men earn because they say they cannot manage it properly. However, our culture is to ask men for permission*.” **A woman from El-Laheley**
>
> “*Income is under the control of women. We are the ones who know everything about what the family needs*.” **A woman from Gad**
>
> “*Women can join any group and do not need permission. The family benefits, and men are positive about it*.” **A woman from Dhaga Jabis**

Most participants explained that women had recently experienced a sense of empowerment in accessing credit, while men seemed to face challenges in gaining creditors’ trust. A woman from Guyo further emphasised this change in credit access with her insights into gender roles and responsibilities.

> “*Women are trusted, while men are not reliable. Creditors place their trust in women. In recent years, there has not been a single responsibility that women could not shoulder. Men used to have far more tasks and greater responsibilities.*” **A woman from Guyo**

Most participants expressed that they have control over expenses and purchasing decisions, attributing this responsibility to women’s oversight of family needs while still acknowledging men’s role as the ultimate decision-makers.

“Women are the ones who make decisions because men do not think about what is needed, such as live animals, food, and clothing.” **A woman from Gad**

> “*Women know what is missing and what is needed. Men just need to know and agree*.” **A woman from Biyo-Didley**

Participants expressed a sense of gender equality in access to training opportunities and groups while also recognising a shift over time. Some women recounted feeling marginalised in the past, though they did not specify whether this was due to being overlooked by training providers or because they did not view animal health as part of their domain. Additionally, when discussing access to groups, women emphasised the benefits for the family.

> “*Previously, women were feeling that they were lower than men. However, now we are equal in terms of participating in training*.” **A woman from Gad**
>
> “*Women can join any group and do not need permission. The family benefits, and men are positive about it*.” **A woman from Dhaga Jabis**

Women expressed a sense of self-determination in managing their time, particularly those involved in income generation. However, the prevailing sentiment among most women was that their workload is dictated by the combination of reproductive and productive tasks they are obligated to fulfill for their family’s sustenance.

> “*Mothers make decisions about their workload. If the woman makes an income, she is responsible for her daily workload. However, she has no choice; life dictates it*.” **A woman from Gad**
>
> “*Men are the rule makers, but women take the lead when it comes to being productive. Women never sit at home; they are always working. Some sell milk, while others sell khat. A man is often just the eldest child. Although some work, the majority do not*.” **A woman from Garbi**

##### Subtheme: Cultural norms and masculinities

The second subtheme, ‘Cultural norms and masculinities,’ delves deeper into women’s perceptions of men as hierarchically superior, possessing final decision-making authority, and undertaking traditional male tasks. Women generally viewed men as the primary owners of livestock and as protectors of women and children, often consulting their husbands for decision-making out of respect. Tasks requiring physical strength were typically associated with men’s responsibilities. Additionally, khat chewing emerged as a social activity among men that was deeply embedded in the culture.

All participants agreed that men are perceived as holding ultimate decision-making authority and ownership of livestock. It was commonly understood that women would seek permission if their husbands were present, with most women indicating that they sought approval from their husbands as a gesture of respect.

> “*Except for the animals we were given for the engagement, the decision about our livestock is not ours to make. As a woman, you should consult your husband about everything. Otherwise, he may not feel respected, weakening trust and potentially leading to divorce*.” **A woman from Biyo-Didley**
>
> “*Women take care of the animals, but men are the owners of the animals*.” **A woman from Garbi**

Some participants identified tasks commonly associated with men. These tasks were characterised by physical demands and potential hazards and were typically perceived as male-dominated responsibilities.

> “*Regarding livestock nutrition, women are responsible for feeding, watering, grazing, and milking. Camels fall under the responsibility of men. Men also handle fencing for livestock, work on the farm, and engage in construction and digging*.” **A woman from El-Laheley**
>
> “*Women and children take the livestock out to graze. Men take on this task during famine due to the far distances involved*.” **A woman from Gilla**
>
> “*Men handle the heavy tasks that women may not be strong enough for, such as construction*.” **A woman from Garbi**
>
> “*He is responsible for protecting the children and me, and I care for his belongings*.” **A woman from Guyo**

Participants elaborated on how men avoided traditionally female tasks, leaving women to handle household chores. Men seemed to engage in khat chewing most of the time and were criticised for not providing adequate support to women. However, the participants seemed to recognise and accept this situation.

> “*Cultures do not allow men to do what women do*.” **A woman from Gad**
>
> “*Most of the time, we blame men, even if he is sleeping; I blame my husband because I do most of the work. We criticize our husbands because we feel stressed when we lack support from them. However, we follow our religion, which emphasises mutual respect between husband and wife. Without respect, there can be no family. Women are expected to respect men because they hold a superior position in the family*.” **A woman from Biyo-Kabobe**

##### Subtheme: Men losing track

Women sometimes sought permission from a “sleeping man,” as men often slept due to khat consumption. This behavior led many men to avoid their responsibilities or become unable to work. This phenomenon is explored in the third subtheme: ‘Men losing track.’

Global and local changes seem to have contributed to a high prevalence of khat consumption, altering lives. According to participants, as men relinquished more responsibilities and obligations, women took on increasingly more of these roles.

> “*Men used to do much work. They were knowledgeable about every aspect of community and household affairs. About 90% of tasks were handled by men, while women primarily focused on cooking. I simply seek his permission now, but he is unaware of what is happening! With the emergence of khat, men have become lazy. Even if he receives some money from family members, he hands it to the women, and I take care of it. We must seek permission, sometimes even from a sleeping man. For men, there are only two responsibilities left: making decisions and handling camels. However, women oversee everything, and they are aware of all household activities. If he brings money, it barely lasts for four days. I am the one who can effectively manage the household. And the livestock. Women take the livestock to pastures because men lack knowledge about livestock and the care they require. He spends his time chewing khat and sleeping*” **A woman from Biyo-Kabobe**
>
> “*Women take care of the livestock. Men are unaware of their needs. We, along with our accompanying children, are the ones who can identify issues with the livestock or recognise when vaccination or treatment is necessary. Sometimes, even when I inform him that the livestock requires vaccinations, he simply walks away*.” **A woman from Gilla**

Participants reported that men were often absent for most of the day due to khat consumption, with some even being unable to work or attend meetings.

> “*Women handle everything. No one trusts men because they chew khat. Women bear the responsibility. Men who chew khat do not work. They chew khat all night and sleep all day*.” **A woman from Biyo-Kabobe**
>
> “*Men may miss meetings; they sleep most of the time because they chew khat*.” **A woman from Biyo-Didley**

Men also resorted to selling livestock to buy khat, potentially creating financial problems for the family.

> “*In every household, men chew khat without exception. My husband gains money from selling the livestock. He selects the finest livestock and sells them for khat. These livestock could have provided security for us, perhaps for my child’s marriage. He dismisses it if I suggest not selling certain cattle, claiming it is his business alone. Some men chew khat day and night*.” **A woman from Gilla**
>
> “*The men forget about their wives and children. Their world revolves solely around khat. They forget about their family’s needs*.” **A woman from Guyo**

Despite expressing challenges with the changes khat introduced and hoping for a return to normalcy, participants expressed acceptance of the current circumstances.

> “*We hope to share the tasks, or at least he brings in the income. We do not like that our husbands chew khat, but we have adapted. I do not criticize it. Everything I have, I give to him, just like I give to my children. He is like my child. However, we cannot live without men. When there are no men, there is no life, no safety. What he does may seem insignificant, but we cannot live without him. And the children listen to their father*.” **A woman from Guyo**

Some women cultivated and/or sold khat to generate income, recognising its economic benefits. However, men were seen as losing track and neglecting their responsibilities due to their khat consumption.

> “*We experience both gains and losses from khat. Khat is useful because it provides support; we sell it, earn income from it, and have money. On the other hand, it is a loss because our husbands have lost their way by chasing after khat and forgetting their responsibilities and their families. Men who do not chew khat are not on the same level as others because those who resist chewing khat help their family and understand their family’s situation. However, most chew khat and are solely focused on obtaining it*.” **A woman from Garbi**

#### 3.1.2 Knowledge privilege, dilemmas, and gaps

This theme delves into the diverse experiences of women living in communities with and without prior PPR training. It examines the empowerment experienced by women who received PPR training – referred to as ‘vaccinators’ in the following sections – as well as the empowerment sought by other participants through animal health training. It also addresses the ongoing dilemmas and challenges faced by communities due to training gaps. We identified three subthemes: ‘Knowledge for empowerment,’ ‘Dilemmas despite knowledge,’ and ‘Challenges due to training gaps.’

##### Subtheme: Knowledge for empowerment

The subtheme ‘Knowledge for empowerment’ explores the experiences of women who reported that animal health training enhanced their knowledge and skills, thereby empowering them. Most participants from communities with PPR training, particularly the vaccinators, reported experiencing a heightened sense of competence, strengthening their confidence and self-efficacy in managing animal health.

For example, a vaccinator from Biyo-Didley described her transformation from a state of “not knowing” to becoming “knowledgeable” and self-sufficient. Furthermore, livestock assets were recognised as invaluable resources.

> “*We know PPR, the symptoms are lesions of mucous membranes, nasal discharge, diarrhea, dehydration, abortion. We treat our livestock ourselves. Vaccination requires experience and should be done by trained women. We also go to the surrounding villages and do the vaccinations. It is a big change because we did not know anything before this training. Now, we are respected because we have learned something; we are knowledgeable. Since animals are the most important asset in the community, training in AH is very important because you must take care of the animals just like you care for the people. In the past, livestock used to die from PPR. Now, we go to the animal health post / private veterinary pharmacies and get medicine, or there is a vaccine that we can administer to protect our livestock. If we have access to the vaccine, the animals do not get sick as much and do not die as often*.” **A woman (vaccinator) from Biyo-Didley**

Most participants acknowledged vaccination as a highly effective method for protecting their livestock and expressed an understanding of its benefits. Specifically, women from communities with PPR training expressed a preference for vaccination over curative treatment, though not exclusively. A woman from Biyo-Kabobe, a village neighboring a community with PPR training, underscored this point:

> “*If we get the vaccinations, we do not see PPR until 6 months. Most goats and sheep will die if we do not get the vaccination. There is nothing better than vaccination. In our village, we never had training. Sometimes, we try other medicines, but they are of no use. Whenever we need vaccines, we go to the other village*.” **A woman from Biyo-Kabobe**

Despite the overall positive perception of vaccination, one participant (a vaccinator) recognised the existence of vaccine mistrust in other communities and expressed a sense of empowerment in advocating for vaccination.

> “*Part of the population still believes that vaccination is used to reduce livestock. This causes them to resist. Nevertheless, we can see that a lack of knowledge causes this problem. If you sit with them and discuss, you can create acceptance*.” **A woman (vaccinator) from Biyo-Didley**

Participants from most communities with PPR training reported that vaccinators are respected within their community and feel welcomed by livestock keepers who seek their services for livestock vaccination. This sentiment was echoed by a vaccinator from Biyo-didley, who emphasised the role of a supportive chairman in fostering this environment.

> “*We are welcomed and treated with respect. The community supports us by catching the goats and organizing the program. Regarding vaccinating at the community level, we receive support from the chairman of the Kebele, as there are water points where many livestock gather. The village chairman mobilizes other communities. Information can be exchanged at the water points, and livestock is collected*.” **A woman (vaccinator) from Biyo-Didley**

In veterinary services and animal health training, women seemed to encounter an environment where they experienced a sense of equality.

> “*Nowadays, women decide if they want to participate in [animal health] training; that is their decision. And if they do not, it is their decision*.” **A woman from Gad**
>
> “*Yes, we go directly to private veterinary pharmacies. Whoever goes will have access; there is no problem. We can even order by phone. As long as you have money, there is no difference between men and women. We are not treated disrespectfully because we are women. It is like a store. Disrespect happens but is not common in animal health*.” **A woman from Dhaga Jabis**

Most participants expressed a strong interest in animal health training, emphasising the vital role of livestock health in sustaining their livelihoods while also anticipating potential business opportunities.

> “*We need to learn more about animal health and educate ourselves on livestock diseases and their clinical signs because livestock production is essential to our livelihoods*.” **A woman from El-Laheley**
>
> “*We want to know the diseases of our livestock. We want to be able to protect our livestock. We want to have access to training. Then we would be able to participate in animal health services*.” **A woman from Gad**

Women who had not received PPR training viewed vaccinators as role models. Vaccinators were not only respected for their expertise but also inspired a strong interest among other women to pursue training in animal health.

> “*It is amazing what these vaccinators are doing. They even vaccinate camels. Many become rich and independent; the vaccinators can travel anywhere and have access to vaccines and cold chains. I hope we will be given the same opportunity.*” **A woman from Biyo-Kabobe**

In every FGD, women expressed great enthusiasm for the ‘train the trainer’ approach and embraced the opportunity to learn from other women. Participants underscored how their empowerment is driven by acquiring knowledge that benefits both themselves and their communities.

> “*We are all prepared. When the time comes, we will participate in animal health training. We would greatly appreciate the opportunity to receive training from another educated woman from the village who can share her knowledge. We aim to attain a high level of animal health. We will join hands and teach each other. It would be beneficial if someone could come into the community and provide training to all interested. The objective is to increase the number of people capable of administering vaccines. Women need to participate in the training. Women can then share the information with other women and men. In the past, men were trained, but they are not active now because they are busy chewing khat! When you train a woman, you train the whole community*.” **A woman from Biyo-Didley**

##### Subtheme: Dilemmas despite knowledge

The subtheme ‘Dilemmas despite knowledge’ explores vaccinators’ challenges after participating in PPR training. Some shared experiences of being rejected by livestock keepers for various reasons. Similarly, obtaining the vaccine during times of need presented a significant challenge, especially in the dry season when resources are scarce at all levels, leading to depleted vaccine stocks and a shortage of ice. Additionally, participants mentioned how safety concerns restricted their mobility, posing another obstacle to accessing or delivering vaccines.

A vaccinator from Biyo-Didley recounted her experience with rejection based on gender, possibly stemming from a lack of trust in women’s competency due to the historical predominance of males in animal health services, as articulated in the second part of the statement.

> “*We may encounter a man who already knows us and will not accept us to administer vaccines to his livestock for personal reasons. Previously, a known man typically performed animal health services, such as a community animal health worker. The community may doubt the vaccinator’s abilities because their training was shorter than the community animal health workers. However, this is only rarely the case.*” **A woman (vaccinator) from Biyo-Didley**

In El-Laheley, a woman explained that rejection in her community was driven by economic factors. Livestock keepers were reluctant to compensate women vaccinators for their services and vaccines, as they were accustomed to receiving free vaccinations for their livestock from the government.

> “*Some of us received training but did not administer vaccines because they expected to be paid for their services. However, the government intervened and provided free vaccinations.*” **A woman from El-Laheley**

Another potential challenge seemed to arise from the community’s reluctance to pay for services provided by women due to gender reasons. This perspective was expressed by a woman who participated in animal health training (though not in the PPR training).

> “*In our community, certain tasks are exclusively reserved for men due to community beliefs. When men perform these tasks, they typically receive a salary. However, women do not receive any payment when they undertake the same work. I vaccinated numerous goats and sheep without receiving any compensation from the community. In our village, women are not remunerated for their efforts. After completing the training, I received equipment I used during the dry season. However, I had no money left to purchase vaccines afterward*.” **A woman from Gilla**

A few participants mentioned that certain communities hold the belief that vaccines cause abortions. A vaccinator from Biyo-Didley explained her approach to addressing vaccine hesitancy in the following statement:

> “*Communities have different views. Some will tell you that vaccination can cause abortions. Then, we sit down and educate them about the benefits of vaccination. First, we explain to them the type of disease, its effects, and the effect of the vaccine. Then, when they understand, we start vaccinating. Some of the community will still resist. We leave their livestock.*” **A woman (vaccinator) from Biyo-Didley**

Furthermore, the limitations or unavailability of vaccines, along with challenges in maintaining a cold chain, posed significant difficulties – especially in areas with high temperatures during the dry season. Resources became scarce due to famine, and livestock diseases became more prevalent.

> “*The famine and PPR go hand in hand, aiding each other, which is why livestock dies*.” **A woman from Gilla**
>
> “*PPR vaccines are only available at private veterinary pharmacies, where they are sometimes not available due to depleted supplies. We have small vaccine boxes that can be used for one day. The challenge for the vaccinators is obtaining ice.*” **A woman (vaccinator) from Biyo-Didley**

Some participants from communities without PPR training identified cultural norms or safety issues as the primary barriers preventing them from leaving their community. Consequently, they anticipated that this restriction would limit their participation in training activities outside the village and hinder them from delivering vaccines to neighboring villages.

> “*As a mother, it would not be an issue for me if my daughter leaves the community to participate in training or deliver vaccines. However, even if I agree, her father would not allow it. A father would never permit his daughter to go to another village where she would spend the night. Since she is a girl, we are unsure what might happen to her in another village. Our culture believes education or work should be done solely within the community. However, it would not be a problem within the community itself.*” **A woman from Biyo-Kabobe**

##### Subtheme: Challenges due to training gaps

The subtheme of ‘Challenges due to training gaps’ addresses two key issues. Firstly, it explores the challenges reported by participants from communities without PPR training, who expressed feelings of being uninformed and dependent. Secondly, it examines challenges arising from the lack of training, including mistrust in vaccines and the overuse of antibiotics.

Most participants from communities without PPR training expressed a sense of being uninformed, suggesting potential gaps in communication, education, or access to information within these communities.

> “*The government has vaccinated our livestock. However, we do not know why or against which diseases the government administers vaccines*.” **A woman from Guyo**

The sense of dependence manifested in two ways. Firstly, participants from communities without PPR training felt dependent on the villages where vaccinators were trained, a situation they preferred to avoid. Secondly, a participant from El-Laheley expressed reliance on the government, which led to a feeling of helplessness when the response was delayed.

> “*We do not have vaccinators, and even if we wanted to, we do not have access to vaccines. If we were trained, it would be easier because we could administer them ourselves and would not need to beg others. When I ask them to provide me with the vaccines so I can administer it myself, the vaccinators from the neighboring village refuse, saying that I do not know how to do it. However, they administer the vaccines for us when we bring the cattle to their village.*” **A woman from Biyo-Kabobe**
>
> “*Due to a lack of accessibility and communication, we may miss out on animal services. Whenever there is an outbreak, and we submit a notification or request, the Pastoral Development Office takes a long time to respond. Since it is beyond our control, the response time can be lengthy. This delay could result in the death of most of the livestock.*” **A woman from El-Laheley**

Some participants from communities not involved in any animal health training expressed the belief that vaccination causes abortion. Despite some voices raised against this mistrust in the same discussion, it seemed to be a subtle concern that may not have fully surfaced.

> “*Whenever my livestock is vaccinated, they do not give birth. Vaccinations cause abortions. Last year when our livestock were vaccinated, they did not even get pregnant. I dislike it when people vaccinate because I know that something bad is going to happen whenever they do.*” **A woman from Gilla**

While administering vaccines was recognised as requiring expertise, and access to vaccines is restricted to trained vaccinators, many participants expressed confidence in administering “injections” (antibiotics) to their livestock. In contrast to the unreliability of vaccine availability, antibiotics appeared to be easily and consistently accessible.

> “*We treat our livestock ourselves. Most women in the community know how to administer injections; however, vaccination requires expertise and should be carried out by trained individuals.*” **A woman from Biyo-Didley**
>
> “*Vaccines are not always available, while treatment is always available.*” **A woman from Biyo-Kabobe**

#### 3.2 Key informant questionnaires on division of labor and women’s empowerment

##### Division of labor in livestock management

The division of labor collected from the twelve KIQs was species-specific, with men primarily responsible for camels and cattle, while women were equally involved in caring for small ruminants (Figure 2). Notably, the figure highlights a gendered division of tasks, particularly in medical treatment (healthcare), which was predominantly handled by men for all types of livestock, according to the KIs.

**Figure 2:**
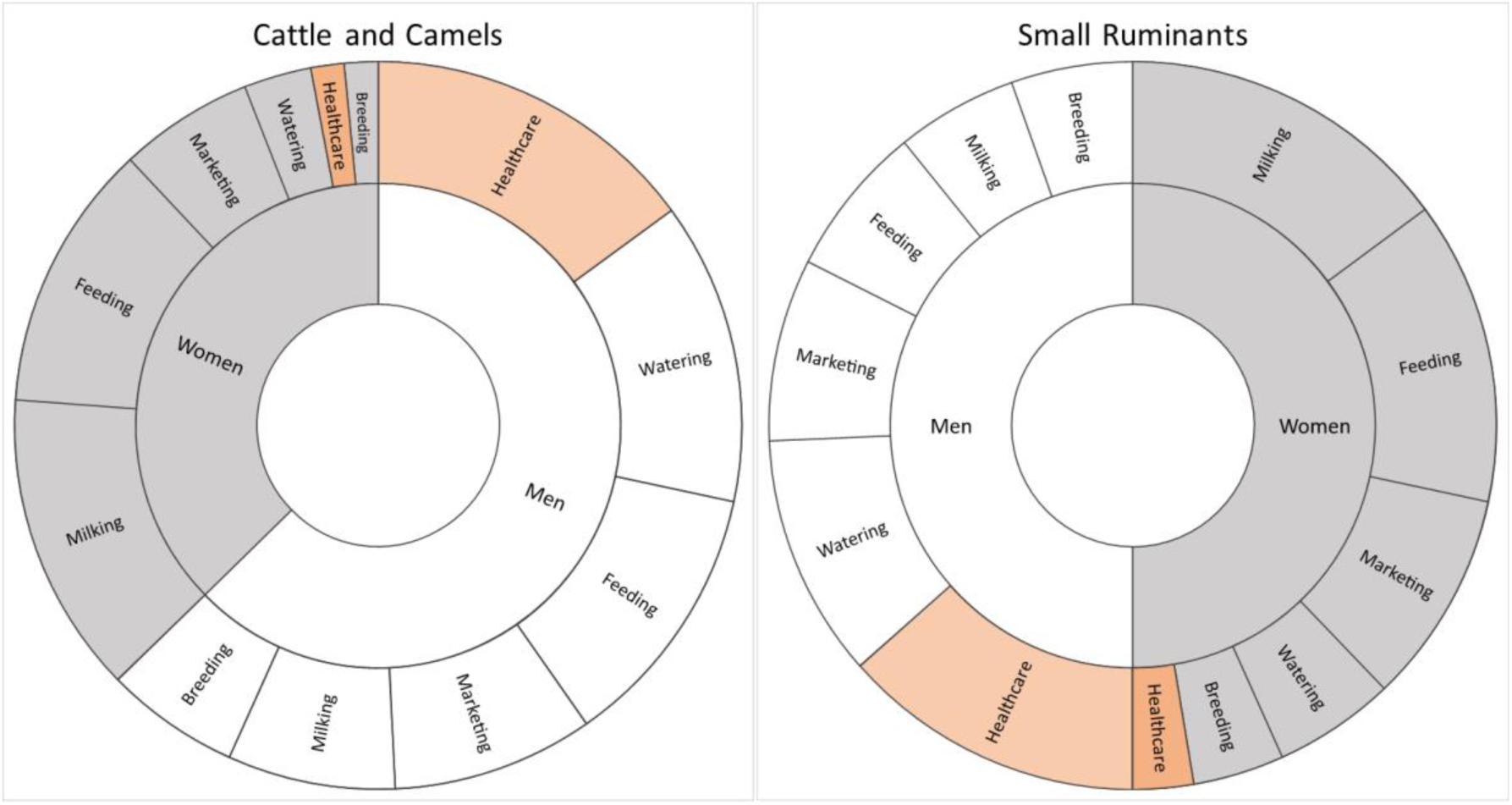
Sunburst diagram illustrating the distribution of key informants’ responses to multiple choice questions on the gendered division of labor related to small ruminants and cattle/camels. “Healthcare” is highlighted in orange for better visibility.

One KI elaborated: *“Livestock-keeping household members, including men, women, and children, have specific and overlapping roles in livestock management. Men provide overall guidance; they identify grazing fields, direct large animals like camels and cattle, milk lactating camels, ensure herd health, and sell large animals. They also construct livestock kraals and accompany cattle and camels for long-distance watering. Women, on the other hand, take care of calves, clean kraals, maintain milking equipment, and sell milk. Children primarily engage in herding, feeding, watering, and assisting in controlling and counting livestock.”*

##### Challenges to women’s empowerment in animal health

KIs acknowledged various obstacles to women’s empowerment in livestock management and animal health in the Somali region of Ethiopia. Nine KIs highlighted the lack of education and training opportunities for women as a significant barrier. Additionally, nine KIs recognised the challenge posed by gender norms in the division of labor. Finally, eight KIs noted the limited access of women to resources and services.

One KI provided a nuanced perspective: *“There is not a barrier; rather, it reflects the division of labor. There are fewer female animal health workers compared to males because girls prioritise other professions. Women hold high respect in the Somali community. Consequently, they are not involved in labor-intensive or dangerous activities. Traditional healers are mostly men.”*

KI’s responses to all questions of the KIQ are available in the supplementary materials (S1_Table).

### 3.3 Key informant interview on men’s consumption of khat

Although khat has existed for decades, its accessibility was initially limited by transportation challenges. However, over the past 30 years, improvements in transportation services have made it more widely available throughout the region. According to the KI, khat consumption was common among both men and women in urban areas, while in rural villages, it was predominantly consumed by men, including young males.

> *“90% of the population has this problem, and most people are busy chewing khat. When the track comes, life comes. It started with the fathers first, then reached the youth, and now both the youth and the fathers are chewing it. The issues got out of control. There are two types of khat chewers: Those who consume excessive amounts reach a point where it drives them into madness. The more they consume, the greater the impact on their mental state, and they find it difficult to quit or reduce their khat intake. Consequently, they cannot work as they sleep during the day. For those who consume small amounts, khat does not adversely affect their mental state, and they can quit whenever they choose. They can work and fulfill their family responsibilities. From an economic perspective, however, they are identical. They destroy much money*.” **KI (PDO)**

The challenges associated with khat seemed diverse: household finances were strained as individuals spent all their money and assets on buying khat, making it difficult for families to afford their children’s education. The inability of khat consumers to work also negatively impacted the broader economy.

> “*People waiting for khat become blind to everything else and often start fighting. However, when khat arrives, and they consume the first leaf, they feel better and calm*.” **KI (PDO)**

For some, particularly women, khat provided an income-generating opportunity.

> “*It has become the most productive and beneficial work in terms of efficiency. People transport it from one place to another, and selling takes less than an hour*.” **KI (PDO)**

The government was identified as the only entity capable of addressing this issue. However, it appeared reluctant to take action, likely because it benefits from the khat trade, with a substantial portion of its revenue coming from khat taxes.

> “*The only entity that has the solution to this problem is the government. The government should have controlled it. It must discipline the citizens; if they do that, they can easily stop this problem. But no, because the government itself benefits from this, as most of the government’s tax revenue comes from khat*.” **KI (PDO)**

## 4. Discussion

This research positions itself within the collaborative efforts of diverse researchers across various contexts, locations, and timeframes to achieve a comprehensive understanding of the topic. Detailed descriptions of the study’s context, participants, settings, and circumstances are provided to enhance the potential for transferability of the results and subsequent discussion. In this context, transferability refers to qualitative research that is deeply contextualized, enabling readers to evaluate the applicability of the analysis to their own settings (Braun & Clarke, 2021).

### 4.1 Navigating hierarchies and the feminisation of responsibility

The first theme, ‘Navigating hierarchies and the feminisation of responsibility,’ explores the importance of pastoral women’s sovereignty in PPR vaccination, given the social context of their livestock-based livelihoods. Participants reported a shift in gender roles, with women increasingly taking on a more prominent role in decision-making processes. This shift is consistent with previous findings among Tanzanian women (Galiè et al., 2019). However, recent research by Haug et al. (2021) highlights the persistence of male dominance in decision-making in Tanzanian and Ethiopian contexts, in contrast to countries such as Malawi, Rwanda, and South Africa, where women frequently take the lead in agricultural decision-making. These contrasting findings point to a dynamic and evolving situation, underscoring the complexity of evaluating decision-making processes across different regions. In Nepal, a similar shift has been observed, where the feminisation of agricultural labor has evolved into women gaining control over decision-making in wheat farming (Farnworth et al., 2019). Farnworth et al. (2019) attribute this shift to several factors, including male outmigration, NGO initiatives promoting gender equality, increased confidence among women, and “strong cooperation among women to support each other’s innovation journeys.”

On the other hand, weak institutional recognition of women as primary livestock keepers continues to create barriers. According to Farnworth et al. (2019), agricultural institutions often fail to acknowledge changing gender dynamics in livestock management, perceiving women solely as helpers, which limits their access to training, finance, and innovation. This perception bias is evident in the disparity between the KIs’ perceptions and the experiences of the female focus group participants in our study. Such bias can lead to unequal targeting of livestock development activities (Kariuki et al., 2022), marginalising women, and neglecting their livestock-related needs (Bryan et al., 2018; A. Quisumbing et al., 2019). However, emerging evidence, including testimonials from our focus group participants and NGO reports, suggests that women are now gaining better access to animal health services, training, and credit opportunities in Ethiopia.

Galiè et al. (2022) advocate for empowering women in livestock management by recognising them as livestock keepers and improving their access to resources, opportunities, and decision-making roles. Most of our KIs agreed with these recommendations, and our focus group participants expressed feeling empowered in these areas. However, our findings indicate that while progress is being made, these measures alone are insufficient to genuinely empower women through livestock interventions. Persistent gender inequalities remain in terms of workload, family and livestock responsibilities, mobility (due to safety concerns), and control over their own time. As Haug et al. (2021) suggest, relying solely on decision-making and self-efficacy as indicators of women’s empowerment within livestock interventions may overlook broader empowerment aspects, particularly regarding well-being outcomes.

Farnworth et al. (2019) highlight the substantial labor demands on women, who balance household and caregiving duties, farm responsibilities, and micro-business endeavors. Our findings support this narrative, revealing that women increasingly take on traditionally male roles in rural households, such as managing livestock marketing and animal health, while still shouldering the majority of reproductive labor. Chant (2014) similarly observes that women of all ages engage in income-generating activities while bearing the majority of reproductive tasks for male household members, increasingly bearing the burden of poverty management and household sustenance. This trend, termed the ‘feminisation of responsibility and obligations,’ is becoming more pronounced as men avoid the everyday demands of domestic life, often engaging in escapist behaviors such as substance abuse (Chant, 2014; Farnworth et al., 2023). In our context, khat consumption has contributed to this shift, leading to job losses and a transfer of responsibilities to women. At the same time, khat offers an income opportunity, particularly for women. However, despite women taking on a growing share of responsibilities in addressing poverty, there has been no corresponding advancement in their rights. Men continue to retain traditional privileges, such as livestock ownership and decision-making authority. Further research is needed to explore the complex effects of khat on social dynamics in rural households and to better understand the perspectives of both women and men regarding these shifting roles.

### 4.1 Knowledge privilege, dilemmas, and gaps

The second theme, ‘Knowledge privilege, dilemmas, and gaps,’ explores the empowering aspects of women’s sovereignty in PPR vaccination while identifying obstacles to achieving full sovereignty.

Knowledge privilege is evident in the disparities we found in both PPR knowledge and agency. Participants from communities with prior PPR training expressed confidence in identifying and managing the disease, while participants from communities without prior training felt inadequate and helpless in dealing with livestock diseases.

While many women who participated in the PPR training felt empowered, two distinct dilemmas emerged: the first involved scenarios where vaccinators were confident, and livestock keepers welcomed their services, but vaccines were unavailable. Unlike findings from Northern Ghana, where vaccine distribution favored men (Omondi et al., 2022), our data suggest that women do not face challenges in accessing vaccines due to their gender. However, vaccine supply chains are complex, involving various actors and activities at different levels, progressing from national to local communities (McKune, Serra, & Toure, 2021). Bottlenecks in production can cause delays in distribution, depleting stocks at animal health posts and/or private veterinary pharmacies (McKune, Serra, & Toure, 2021). Building upon the approach taken by McKune et al., who conducted their research in Senegal, we recommend a gender-sensitive analysis of the vaccine supply chain and distribution system in the Somali region to understand how gender-related factors influence access to vaccines and participation in the system. Such an investigation would lay the groundwork for future PPR intervention projects.

The second dilemma involved livestock keepers rejecting vaccinations offered by vaccinators due to personal reasons, such as gender biases, costs, or mistrust of vaccinations.

While many women, particularly those from communities with PPR training, affirmed their ability to perform tasks traditionally associated with men, some reported having encountered doubts about female competence. This situation reflects entrenched gender norms that restrict women’s roles, with some questioning whether it is culturally acceptable for women to perform tasks traditionally assigned to men (McKune, Serra, & Toure, 2021). Nonetheless, women from communities with PPR training expressed confidence in managing livestock, and vaccinators were seen as role models, inspiring others to seek similar opportunities. This finding highlights an additional facet of the empowering impact of PPR training: the presence of role models can effectively inspire and reinforce individuals’ belief in their capacity for change (Eger et al., 2018).

Rejections may also stem from livestock keepers’ unwillingness to pay for vaccines, which is a primary barrier to vaccine uptake in study sites in Uganda and Kenya (Mutua et al., 2019). This reluctance may arise from the belief that livestock keepers should not incur this cost when vaccines are provided free of charge by the government.

Another reason vaccinators face rejection is skepticism about vaccine safety. The belief that vaccines cause abortions was expressed exclusively by participants from communities without prior PPR training. This skepticism may stem from negative experiences with vaccines such as the Smithburn vaccine for Rift Valley fever, which has been associated with adverse effects (Kortekaas et al., 2012). Such experiences can foster negative perceptions about specific vaccines or vaccines in general within the community (Mutua et al., 2019). However, extensive research on live-attenuated PPR vaccines demonstrates their safety and efficacy (Mahapatra et al., 2020; Milovanović et al., 2023; Saravanan et al., 2010). Therefore, PPR is a disease that is effectively controllable through vaccination, a view widely supported by focus group participants from communities with PPR training. Given the vaccine’s robust safety records, it holds the potential to build greater trust in vaccination practices overall.

Our findings suggest that vaccinators successfully mitigated vaccine hesitancy by sharing vaccination information, which fostered acceptance. Marsh et al. (2016) emphasise the role of other pastoralists and NGOs in enhancing vaccine acceptance and uptake, revealing a notably higher vaccination rate when such entities provide vaccine information compared to instances where animal health services are obtained from commercial vendors. We recommend further studies using a transdisciplinary approach to examine social drivers and barriers to vaccine uptake, involving stakeholders from sociology, human and veterinary medicine, and the vaccine supply chain.

A related concern and challenge stemming from a knowledge and training gap is the potential overuse of antibiotics. Although our data do not quantify antibiotic usage, the availability of antibiotics when vaccines were not accessible, combined with the confidence expressed by focus group participants in administering injections, suggests that this issue warrants further investigation and awareness-raising, especially given the global challenge of antibiotic-resistant bacteria (Woolhouse et al., 2015).

Participants consistently expressed a strong interest in receiving training in animal health and demonstrated a desire to independently manage the health of small ruminants. This includes acquiring skills to diagnose PPR, knowledge about its prevention and treatment, and proficiency in PPR vaccination. The ‘train the trainer’ approach, frequently mentioned during FGDs, emerged as a potential strategy for implementing community-based training. Community-based participatory research and health promotion are fundamental to ensuring the sustainability of interventions (Nutbeam & Muscat, 2021; Pederson et al., 2015). This approach requires active community engagement and empowerment to increase control over the determinants of livestock health. However, further research is needed to assess the feasibility of this approach.

This vision for future initiatives is based on evidence that integrating women’s empowerment with improved vaccine access and knowledge has the potential to significantly increase vaccination rates, improve livestock productivity, and enhance household livelihoods (Omondi et al., 2022).

## 5. Conclusion and recommendations

Efficient vaccine distribution systems targeting livestock diseases managed by women could significantly enhance the livelihoods of female smallholder farmers. However, efforts to increase women’s participation in vaccine administration may further add to their workload, potentially compromising their genuine empowerment. Livestock innovations should carefully assess the trade-offs between the additional time investments required of women and the associated benefits (Dumas et al., 2018; Johnston et al., 2015). According to Galiè et al. (2022), the question is wether livestock interventions should support women’s livelihoods (A) by easing their role as caretakers or (B) by offering opportunities to expand women’s livelihoods beyond caregiving roles. This consideration has significant implications for future PPR intervention projects.

For option (A), essential actions include facilitating women’s access to vaccines and cold chain systems through government and/or NGO support and providing widespread community-based training to enhance women’s sovereignty in small ruminant health. This approach promises to reduce time spent on animal health management in the short term while safeguarding the lives of small ruminants and improving livelihood security in the long term, which in turn further reduces the workload over time. For option (B), additional actions include offering training in financial management and facilitating access to finance, enabling women to create new income opportunities, such as starting businesses in animal health services. This option would also require empowering men in caregiving roles.

Promoting women’s empowerment locally through gender-responsive, transformative approaches is crucial. However, achieving a broader impact requires collaborative efforts from governments, international organizations, civil society, and the private sector to promote gender equality through national policies, campaigns, and large-scale programs (FAO, 2023). This collaborative effort would also provide a strong foundation for sustainably establishing the role of women in animal health services, including in the eradication of PPR.

## Funding statement

This research was funded by the Specialisation Committee (SpezKo) of the Vetsuisse Faculty Bern.

## Conflict of interest declaration

The authors declare no conflict of interest.

## Ethics statement

Ethical review and approval were not required for this study in accordance with Swiss legislation and local institutional requirements. All participants received detailed information regarding the study topic, the discussion’s concept, the estimated duration of the discussion, the use of recording equipment and note-taking, and the management and utilisation of collected data before their involvement in the study. Participants were encouraged to ask questions and were provided with comprehensive answers. Their participation was voluntary, and they provided oral consent before joining the discussion.

## Data Availability

All data produced in the present study are available upon reasonable request to the authors.

## Acknowledgements

We express our sincere gratitude to all the focus group participants and key informants for their invaluable insights and contributions to this study. We especially thank the entire VSF-Suisse team in Switzerland and Ethiopia for their unwavering support and collaboration throughout the research process.

## Author contribution statement

This research was conducted with the contributions of the following authors:

- **Valerie Hungerbühler**: Led the research design, data collection, data analysis, and manuscript writing.
- **Buckary Barkadle**: Coordinated gender-sensitive projects in the Somali region of Ethiopia, facilitated access to study sites, participated in data collection, provided local context insights, and contributed to the manuscript.
- **Amina Mohamed Hussein**: Facilitated focus group discussions, participated in data collection, provided local context insights, and contributed to the manuscript.
- **Sofia C. Zambrano**: Provided expertise in qualitative research methods, assisted in data analysis, and contributed to the manuscript.
- **Mohammed Digale**: Coordinated gender-sensitive projects in the Somali region of Ethiopia, participated in data collection, and provided local context insights.
- **Kebadu Simachew Belay**: Provided project coordination and logistical support, contributed to project organisation and administration, and offered local context insights.
- **Flurina Derungs**: Provided overall organisational oversight, contributed to project administration, offered gender science-related perspectives and knowledge, and contributed to the manuscript.
- **Salome Dürr**: Supervised the entire research process, provided guidance on research design and methodology, and assisted in manuscript writing and revisions.

Each author has significantly contributed to the work reported in this research, in accordance with McNutt et al., Proceedings of the National Academy of Sciences, Feb 2018, 201715374; DOI: 10.1073/pnas.1715374115.

## Data availability

All data underlying the research findings of this study are provided as supplementary material.

## Supplementary material

**S1_Table.** Table summarising the distribution of the 12 KIs’ responses to the following multiple-choice questions.

**S2_Dataset_FGD.** Dataset containing the transcripts of the focus group discussions (FGDs).

**S3_Dataset_KIQ.** Dataset containing the responses to the Key Informant Questionnaires (KIQ).

## Notes

### Competing Interest Statement

The authors have declared no competing interest.

### Author Declarations

Ethics committee of the Gesundheits-, Sozial- und Integrationsdirektion des Kantons Bern waived ethical approval for this work.

